# Risk Stratification of Arrhythmogenic Consequences of Andersen-Tawil Syndrome Affecting Kir2.1-PIP2 Interactions

**DOI:** 10.64898/2026.05.13.26353154

**Authors:** Lilian K. Gutiérrez, Francisco M. Cruz, Álvaro Macías, Ana I. Moreno-Manuel, Patricia Sánchez-Pérez, María Linarejos Vera-Pedrosa, Fernando Martínez de Benito, Aitor Díaz-Agustín, Juan Pablo Ochoa, Juan Manuel Ruiz, Francisco J. Bermudez-Jimenez, Isabel Martínez-Carrascoso, Salvador Arias-Santiago, Aitana Braza-Boïls, Marta Gutiérrez-Rodríguez, Mercedes Martín-Martínez, Esther Zorio, Juan Jiménez-Jaimez, José Jalife

## Abstract

**Background:** Andersen-Tawil syndrome type 1 (ATS1) is caused by loss-of-function mutations in *KCNJ2*, which encodes the inward rectifier K^+^ channel Kir2.1, a key determinant of I_K1_. Impaired Kir2.1 destabilizes membrane excitability and predisposes to ventricular arrhythmias. Most ATS1 variants disrupt channel regulation by phosphatidylinositol 4,5-bisphosphate (PIP2), but whether specific mutations confer differential arrhythmic risk remains unclear.

**Objective:** To determine whether ATS1 variants disrupting Kir2.1–PIP2 interactions define distinct arrhythmic risk profiles and establish a mechanistically informed framework for risk stratification.

**Methods:** We performed a pooled patient-level analysis of 225 ATS1 patients carrying *KCNJ2* variants impairing Kir2.1-PIP2 interaction. Inclusion of 22 clinical and electrocardiographic variables were used to identify mutation-specific risk profiles and predictors for arrhythmia risk. The approach was validated in a multicenter cohort of 20 ATS1 patients. Functional validation was performed using patient-derived iPSC-CMs, cardiac-targeted mouse models, and structural *in silico* analyses.

**Results:** ATS1 variants segregated into three discrete clusters corresponding to high-, intermediate-, and low-risk arrhythmic phenotypes, establishing a mutation-dependent hierarchy of arrhythmic risk. Regression analyses identified six variables independently associated with severe arrhythmic outcomes. In patient-derived Patient-derived iPSC-CM demonstrated graded impairment of electrical propagation and arrhythmia susceptibility, with a hierarchy in CV (Control > R82W > R218W > G215D). ATS1 mouse models reproduced the clinical risk stratification. Structural modeling showed that high-risk variants localize near the channel pore and disrupt Kir2.1–PIP2 interactions through mutation-specific mechanisms.

**Conclusions:** ATS1 caused by Kir2.1–PIP2–disrupting variants is not a uniform disorder but comprises biologically distinct subgroups with predictable differences in arrhythmic severity. Integrating genetics, functional phenotyping, and structural modeling provides a mechanistically grounded framework for ATS1 risk stratification and precision therapy development.

## Introduction

Andersen-Tawil Syndrome type 1 (ATS1) is an autosomal dominant ion channel disease characterized by a triad of clinical manifestations, including periodic skeletal-muscle paralysis, dysmorphic features, and ventricular arrhythmias.^1–3^ Loss-of-function mutations in *KCNJ2*, the gene encoding the strong inward rectifier potassium channel Kir2.1, produce ATS1.^1,4,5^ Kir2.1 dysfunction sets the stage for abnormal cardiac ventricular repolarization, prominent U waves, and QTU interval prolongation.^6^ Most importantly, ATS1 is characterized by significant life-threatening arrhythmias, including bidirectional ventricular tachycardia, polymorphic ventricular tachycardia, cardiac arrest and sudden cardiac death (SCD). In many ATS1 patients, the high burden of ventricular arrhythmias may degenerate into ventricular fibrillation (VF) and ultimately SCD.^7,8^

Kir2.1 channels conduct the cardiac inward rectifier K^+^ current (I_K1_), which stabilizes the resting membrane potential (RMP), controls the threshold for action potential (AP) depolarization, and determines the final phase of AP repolarization.^9^ I_K1_ reduction depolarizes the RMP, which indirectly reduces the cardiac sodium channel (Na_V_1.5) availability by promoting its inactivation, thereby impairing excitability and conduction velocity (CV).^10–12^ Therefore, ATS1 mutations cause a dominant negative effect that interferes with Kir2.1 function and may also affect other ion channels.^13,14^ Importantly, beyond this indirect effect on Na_V_1.5, there may be an additional mutation-specific direct effect on Na_V_1.5, further contributing to the pro-arrhythmia observed in ATS1.^15^

Kir2.x channels are uniquely activated by membrane-delimited phosphatidylinositol-4,5-biphosphate (PI(4,5)P2 or PIP2).^16^ More than 50% of reported variants linked to ATS1 affect Kir2.1-PIP2 interactions.^17,18^ In the Kir2.x protein, specific positively charged amino acid residues form a binding pocket that attracts the negatively charged phosphate groups of the phospholipid and enables Kir2.x-PIP2 interaction.^16^ Gating of Kir2.1 consists of a series of protein structural rearrangements, starting when PIP2 reaches the Kir2.1 binding pocket and culminating with conformational changes that trigger channel activation.^19,20^ Studies on mutations disturbing Kir2.1-PIP2 binding have reported a variety of cardiac electrical manifestations.^18^ Yet, whether different mutations interfering with Kir2.1-PIP2 interactions affect the channel differently to produce arrhythmias with differing degrees of lethality has not been studied systematically.

Since ATS1 was first described in 1971,^2^ many cases with varying symptoms have been reported. Standard treatments include beta-blockers alone or in combination with class Ic antiarrhythmic drugs (AADs), such as flecainide or propafenone. However, these therapies often show limited efficacy, and clinical management becomes challenging, particularly when the underlying substrate is intrinsically proarrhythmogenic due to poorly understood mechanisms. ^7,8,21,22^ Understanding the genetic determinant that modulate arrhythmia risk could therefore provide crucial insights for precision-guide therapy in ATS1. Here we present a study aimed at determining whether certain Kir2.1 gene variants that disrupt PIP2 binding increase arrhythmia risk in a defined subset of ATS1 patients. To address this, we analysed 225 ATS1 cases from the literature to evaluate their arrhythmia risk, including studies that reported individual patient-level data on *KCNJ2* loss-of-function variants affecting Kir2.1–PIP2 interaction and excluding reports lacking sufficient phenotypic or genetic resolution for risk stratification. In parallel, we also conducted functional studies in induced pluripotent stem cell-derived cardiomyocytes (iPSC-CMs) generated from skin biopsies of ATS1 patients, as well as in mouse models carrying selected Kir2.1 mutations known to disrupt Kir2.1-PIP2 interactions. These complementary approaches enabled us to develop a novel mechanistic stratification of ATS1 mutations according to arrhythmia severity, providing a foundation for more personalized and targeted therapeutic strategies in ATS1.

## Methods

### Patient-level pooled analysis for ATS1

A systematic review was performed according to the PRISMA Statement guidelines.^23^ Individual patient data from published reports in PubMed and Web of Science were extracted and pooled for analysis (**Figure 1**). The derivation cohort included male and female subjects with a total of 225 individuals carrying *KCNJ2* loss-of-function mutations that affect Kir2.1-PIP2 interaction. A separate validation cohort of 20 individuals with ATS1 variants was included from hospitals in Spain and the UK. The study was approved by the CNIC and Carlos III Institute ethics committees.

**Figure 1.**
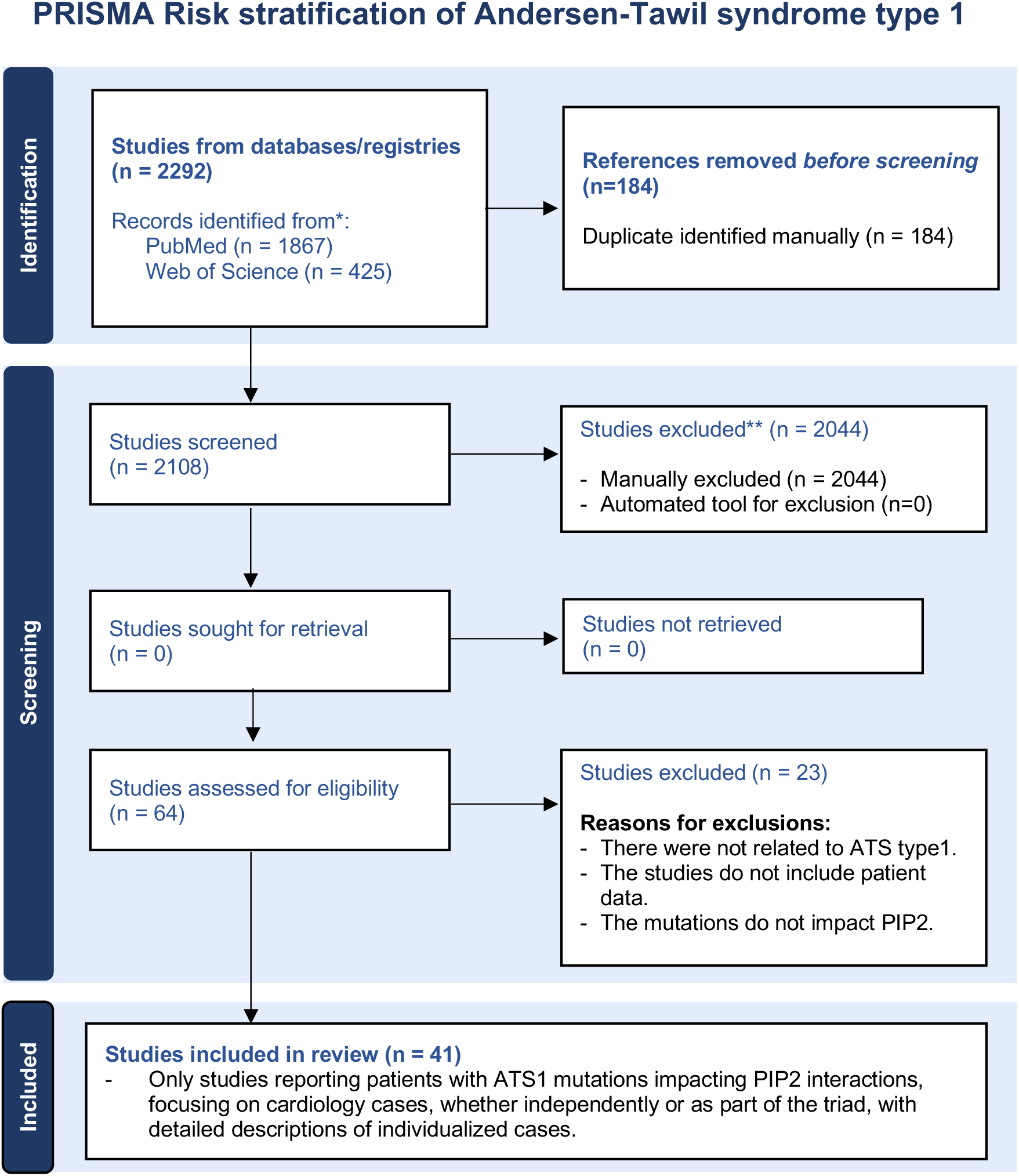
PRISMA flow chart of the screening process. The flow diagram illustrates the selection process. Initially, 2292 studies were identified from two databases. After removing 184 duplicates, 2108 studies were screened for relevance. Of these, 2044 were excluded for not being related to ATS cardiology cases, lacking individual case data, or containing duplicate co-authorships. A total of 64 studies were then assessed for eligibility, with 23 excluded due to inadequate descriptions or insufficient information. Finally, 41 ATS1 studies focusing on cardiology cases with variants impacting Kir2.1-PIP2 interactions were included.

#### Development of Cardiac Risk Stratification

Twenty-two clinical, electrocardiographic, and phenotypic variables associated with ATS1 were analyzed. These variables were selected based on the documented cardiac manifestations within the ATS1 patient cohort and included: sex, history of SCD, ventricular fibrillation (VF), syncope, Torsade de pointes (TdP), polymorphic ventricular tachycardia (VT), VTs, non-sustained VT, bidirectional VT, bigeminy, couplets, trigeminy, VAs, extrasystoles, premature ventricular complexes (PVCs), ventricular ectopy, prominent U waves, right or left bundle branch block (RBBB/LBBB), long QT (LQT), periodic paralysis, and dysmorphic features.

#### Scoring of cardiac manifestations

All the manifestations mentioned above were used for the initial clustering. Selected cardiac manifestations were classified according to severity based on previous studies (**Table 1**).^5,24–26^ Values ranged from 0 (least severe) to 5 (most severe), depending on severity. The Centro Nacional de Investigaciones Cardiovasculares (CNIC) Bioinformatic Unit revised and validated the scoring.

**Table 1.**
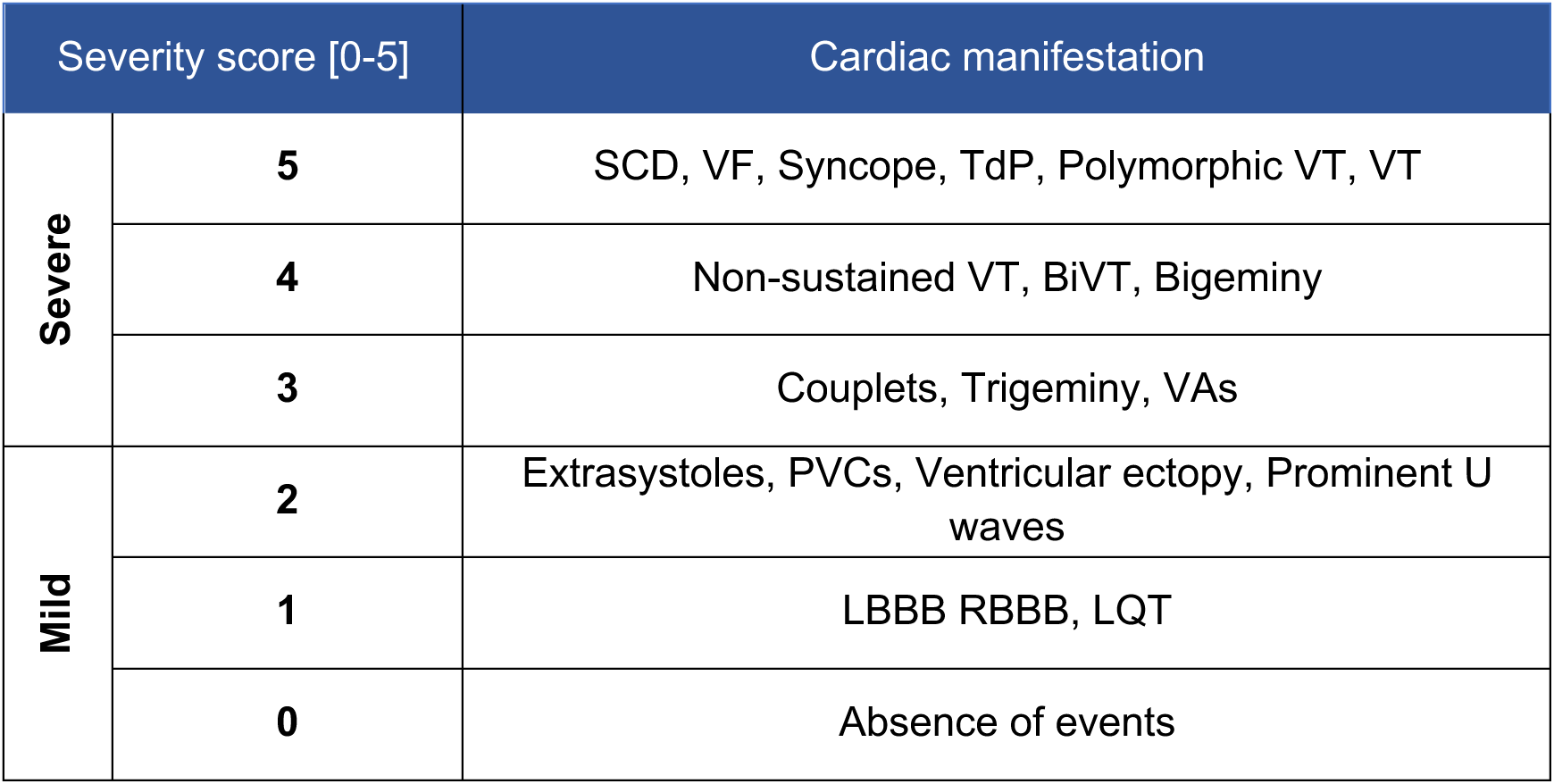
Classification of cardiac manifestations according to severity.

#### Model development

The similarity between variables and patients with different mutations was assessed using hierarchical clustering, including cardiac manifestations, severity score, and ATS1 patients. Clustergram analysis was performed using hierarchical clustering functions of Matlab Statistics and a Machine Learning Toolbox (Mathworks Inc.). Additionally, k-means sub-clustering was performed on hierarchical clustering results to obtain the representative mutations from each group based on the Matlab Statistics and Machine Learning Toolbox (Mathworks Inc.). Stratification of ATS1 variants was obtained by first assigning a severity score to all ATS1 patients ATS1 patient score, as follows:

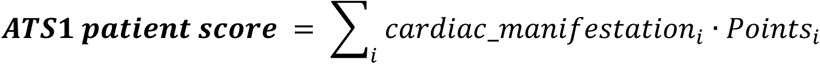

The *mutation score* was computed by gathering all patients for each mutation and calculating a global value for each ATS1 variant according to four distinct statistical measures: mean score, median score, max score, and Jaccard score. Severity score computation and results are performed using Matlab Statistics and Machine Learning Toolbox (Mathworks Inc.).

a) **Mean score**_ATS1 variant_ = 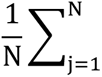 ATS1 patient score_j_
b) **Median score**_ATS1 variant_ = median_j_ ATS1 patient score_j_
c) **Max score**_ATS1 variant_ = max_j_ ATS1 patient score_j_
d) **Jaccard score**_ATS1 variant_ = 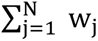 · ATS1 patient score_j_; where weight for each patient is given by, 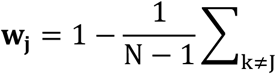 Jaccard ditance ATS1 patient score_j_, ATS1 patient score_k_

#### Model validation

The stratification model of ATS1 variants was validated using data from 20 patients from the multicentric hereditary cardiopathy units in Spain and the UK. Severity score assignment for each ATS1 patient, and *mutation score* were computed as mentioned above in the model development.

#### Regression analysis

Regression analysis was performed using Python libraries *pandas* and *statsmodel*. Data on continuous variables such as age were not used for multivariable analyses because they were not statistically significant. Variables in regression analyses were dichotomized as present (1) or absent (0) in the ATS1 cohort.

### Patient-specific iPSC-CMs

Skin biopsies were collected from three ATS1 patients with different mutations from the validation cohort and one healthy control (Hospital Universitario Virgen de las Nieves, Granada, Spain, and Hospital Universitario La Fe, Valencia, Spain, IRB approvals 2020-582-1 and 2020-411-1). The use of iPSCs and iPSC-CMs was approved by the CNIC Ethics Committee and Madrid authorities. iPSC-CMs were generated and purified after 30 days in culture as previously described (see **Supplemental Material**). Optical mapping was performed after 7 days of maturation as described elsewhere.^27^

### *In vivo* ATS1 mouse model and optical mapping

All animal experiments were conducted in accordance with EU Directive 2010/63/EU, Recommendation 2007/526/EC, and Spanish law (Real Decreto 53/2013). Protocols were approved by local ethics committees and the Comunidad de Madrid (PROEX 111.4/20, 226.5/23). Optical mapping was performed in isolated, Langendorff-perfused hearts loaded with the voltage-sensitive dye Di-4-ANBDQPQ, as previously described.^28^ Cardiac stimulation was conducted using a custom-made bipolar electrode. Conduction velocity (CV) and action potential duration (APD) maps were constructed using trains of 10 pulses (duration, 2 ms; amplitude, 10 V) applied at the ventricular apex at varying basic cycle lengths (BCLs) ranging from 300 to 50 ms in decremental steps of 50 ms, until reaching refractoriness. Arrhythmia inducibility was evaluated using an S1-S2 protocol, using 10 stimuli at 100 ms (S1), followed by 10 stimuli at 40 ms (S2) controlled by a custom-made Matlab user interface, as previously described.^14,29^

### Statistical Analyses

We used GraphPad Prism software versions 8.0 and 10.0 and Python for multivariable and univariable analyses. All included variables were dichotomous. All data sets were tested for normal distribution (Shapiro-Wilk and Kolmogorov-Smirnov tests). In general, experimental comparisons were made using Student’s t-test, as the data were normally distributed. Unless otherwise stated, we used one-way or two-way ANOVA for comparisons among more than two groups and Tukeýs correction for multiple comparisons. Clinical data are expressed as mean±SD, while experimental data are presented as mean±SEM Differences are considered significant when *p<0.05; **p<0.01; ***p<0.001; ****p<0.0001.

## Results

### Novel stratification of ATS1 patients based on cardiac arrhythmic risk

Patient-level pooled analysis comes from 239 individual patients (52.7% female; age 23.3 ±14.8 years) carrying 47 ATS1 mutations reported in 41 published cases between the years 2000 and 2022 in PubMed and Web of Science (**Figure 1** and **Supplementary Table 1**). The search included the keywords: ATS1, Arrhythmias, PIP2, Andersen-Tawil. Patients selected (n=225) were those carrying 39 mutations with PIP2 impairment (**Supplementary Figure 1**).^18^ Characteristics of the cohort are presented in **Table 2**. Among the triad of ATS1 signs, probands included documentation of ventricular arrhythmias (n=147, 65.3%), periodic paralysis (n=110, 48.8%), syncope (n=21, 9.3%), and dysmorphias (n=122, 54.2%).

**Table 2.** Baseline clinical characteristics of ATS1 study cohort (N = 225)

| Baseline characteristics patients |  |
| --- | --- |
| Female, n (%) | 118 (52.7) |
| Age, years | 23.3 ± 14.8 |
| Ventricular arrhythmias, n (%) | 147 (65.3) |
| Periodic paralysis, n (%) | 110 (48.8) |
| Dysmorphic features, n (%) | 122 (54.2) |
| Syncope, n (%) | 21 (9.3) |
| QRS (ms) | 81 ± 9 |
| QTc > 450 ms, n (%) | 42 (18.6) |
| Complete ATS triad, n (%) | 140 (62.2) |
| Medical treatment |  |
| ICD, n (%) | 22 (34.9) |
| βAR-Blockers, n (%) | 14 (22.2) |
| Class I antiarrhythmics, n (%) | 9 (14.3) |
| Acetazolamide, n (%) | 19 (30.2) |
| Values are n (%), age and QRS displayed in mean ± SD. ATS = Andersen-Tawil syndrome, ICD = implantable cardioverter defibrillator. |  |

At baseline evaluation, 62.2% (n=140) of patients presented the complete triad of symptoms, while 37.8% (n=85) showed an incomplete ATS phenotype (**Table 2**). In all cases, medical treatment lacked detail and specificity. The latest European Guidelines recommend three strategies: 1) implantable cardioverter defibrillator (ICD) implantation, 2) beta-blockers, 3) and/or flecainide with or without acetazolamide.^30^ The principal therapies in this study were ICD implantation (34.9%) and beta-blockers (22.2%) for ATS-related cardiac events management (**Table 2**). A fraction of patients (14.3%) was treated with class-I AAD, especially class-Ic drugs (e.g., flecainide). Acetazolamide was reported for periodic paralysis, if present (48.8% of patients).^3^ In accordance with *ClinVar,* 43.5% of the identified Kir2.1-PIP2 variants were considered pathogenic and 15.4% as ‘likely pathogenic’.^31^

Hierarchical clustering was performed to determine whether arrhythmia events could distinguish risk among Kir2.1-PIP2 mutations. Three clusters emerged: #1) mutations associated with a high incidence of severe arrhythmic events (n=150 individuals), #2) mutations linked to mild and heterogeneous cardiac phenotypes (n=43 individuals), and #3) mutations with minor or no cardiac manifestations (n=32 individuals) (**Figure 2A**). Univariate analyses identified key phenotype-specific predictors of ATS1 cardiac severity (**Table 3**). Multivariable regression subsequently selected clinical variables independently associated (p<0.01) with high-risk mutations, based on *KCNJ2* classification, including non-sustained ventricular tachycardia, SCD, bigeminy, and syncope (**Table 3**). Secondary hierarchical clustering restricted to cardiac variables with p<0.05 further improved population stratification, clearly separating severe from mild phenotypes and reinforcing the definition of **cluster 1** (**Figure 2B**).

**Figure 2.**
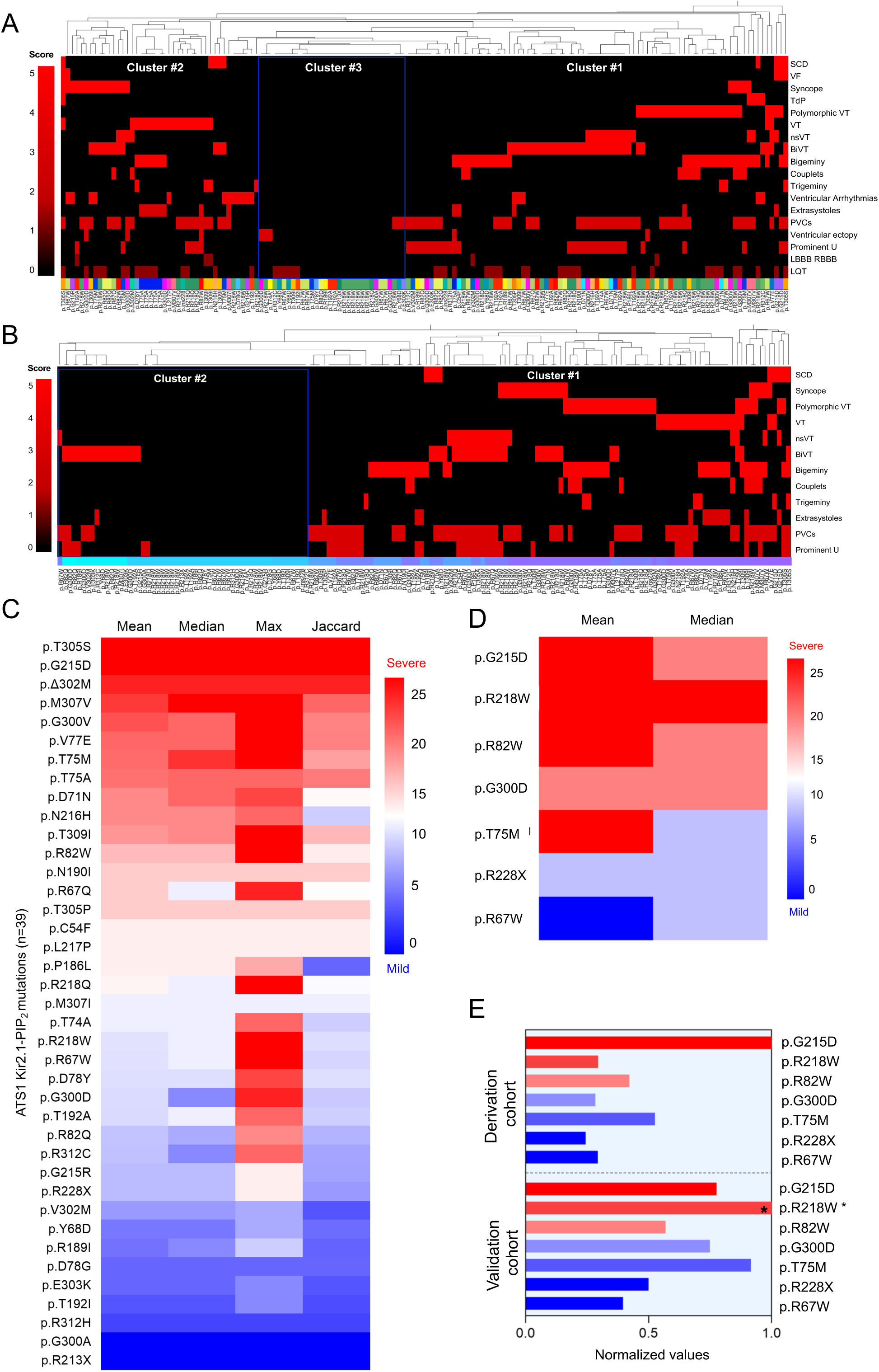
Stratification of arrhythmias in ATS1 mutations. **A.** Hierarchical cluster showing association between mutations and ATS cardiac manifestations (N=225). Three distinguishable clusters result from this classification, being severe (cluster #1), mild phenotypes (cluster #2), and absence (cluster #3). **B.** Restrictive cardiac risk severity discarding non-significant clinical variables yields a more differential phenotype clustering (p < 0.05). **C.** Heatmap of mutations colored by the order of severity determined by each scoring method. High-severity mutations are colored red, and mild severity mutations are colored in blue**. D.** Heatmap showing outcomes of ATS1 cardiac risk stratification of the validation dataset (N=20). Cardiac risk scoring is based on the mean score, while the median score was used for dispersion and reproducibility purposes. **E.** Bar plot presenting the normalized scores to the highest value between the proposed stratification model and the validation dataset. Asterisk (*) indicates a mutation with a single patient.

**Table 3.** Phenotype-specific variables predicting ATS1 cardiac severity.

|  | Univariable Analysis |  | Multivariable Analysis |  |
| --- | --- | --- | --- | --- |
|  | OR (95% CI) | *P* value | OR (95% CI) | *P* value |
| Non-sustained VT | 33.69 (7.35 – 154.47) | <0.0001 | 336.1 (18.4 – 6125.1) | 0.0001 |
| Sudden cardiac death | 10.22 (2.03 – 51.35) | 0.004 | 62.39 (4.65 – 836.91) | 0.0018 |
| Couplets | 9.16 (2.35 – 35.70) | 0.001 | 4.76 (0.81–28.03) | 0.084 |
| Syncope | 6.15 (2.26 – 16.72) | 0.0003 | 33.33 (3.38–328.59) | 0.0027 |
| Trigeminy | 6.93 (1.29 – 37.19) | 0.023 |  |  |
| Bigeminy | 4.32 (2.01 – 9.30) | 0.0001 | 27.41 (3.23–232.78) | 0.0024 |
| Ventricular tachycardia | 3.33 (1.30 – 8.53) | 0.012 |  |  |
| Extrasystoles | 3.28 (1.03 – 10.39) | 0.043 |  |  |
| PVCs | 3.12 (1.52 – 6.39) | 0.001 |  |  |
| Polymorphic VT | 2.87 (1.22 – 6.74) | 0.015 |  |  |
| Bidirectional VT | 2.57 (1.22 – 5.39) | 0.012 |  |  |
| Prominent U wave | 2.19 (1.00 – 4.78) | 0.048 |  |  |
| Age | 1.60 (0.69 – 3.71) | 0.272 |  |  |
| Female | 1.42 (0.71 – 2.83) | 0.327 |  |  |
| Ventricular arrhythmias | 1.43 (0.49 – 4.12) | 0.511 |  |  |
| Long QT | 1.21 (0.58 – 2.54) | 0.611 |  |  |
| Left/Right bundle branch block | 0.62 (0.07 – 5.69) | 0.672 |  |  |
| Ventricular ectopy | 0.83 (0.16 – 4.27) | 0.823 |  |  |
| Torsade de Pointes | - | - |  |  |
| Ventricular fibrillation | - | - |  |  |

### Novel Proarrhythmic Risk Assessment of ATS1 patients with Kir2.1-PIP2 mutations

We applied four quantitative scoring methods including Mean, Median, Max, and Jaccard scores to classify mutations, accounting for phenotypic variability observed among families carrying identical variants. All approaches consistently stratified mutations (**Figure 2C and Supplementary Figure 2A**); however, the Mean score provide the most representative estimate of arrhythmia risk, with high-severity variants clustering at the upper end (red), and low-severity variants at the lower end (blue). Mutations in red indicate the highest pro-arrhythmia risk. This Pro-arrhythmia risk assessment demonstrated excellent performance, with a receiver operating characteristic (ROC) area of 0.93 (p<0.0001; **Supplementary Figure 2B**).

### Clinical Cohort Validates the ATS1 Proarrhythmic Risk Assessment Model

To validate the ATS1 proarrhythmic risk model, the scoring method was tested on an external multicenter cohort of ATS1 patients with Kir2.1-PIP2 interaction mutations. Seven centers in Spain and the UK provided patient data. This cohort comprised 20 patients and represented 17.9% of the studied mutations (n=7 *KCNJ2* variants; **Supplementary Table 2**). Mean scoring effectively identified mutation severity (**Figure 2D**), and the median scoring method was used to assess data dispersion. Both methods aligned with the variant stratification from the broader study (**Figure 2E** and **Supplementary Figure 2C**).

### Patient-specific iPSC-CMs carrying ATS1 mutations present impaired electrical propagation and support proarrhythmic risk assessment

Three patients from the validation cohort (one heterozygous female carrying the *KCNJ2* R82W; one heterozygous male carrying R218W; and a third heterozygous female patient carrying G215D; **Figure 3A**) consented to donate skin biopsies to obtain patient-specific iPSC-CM monolayers. These three mutations covered the range of severity in the stratification of the ATS1 cardiac arrhythmia phenotype. iPSC-CM monolayers from a healthy young male, genetically unrelated to any of the patients were used as control (WT). ECGs and arrhythmic activity were accessible for all three patients, each showing spontaneous ventricular arrhythmias characteristic of ATS1 (**Figure 3B**).

**Figure 3.**
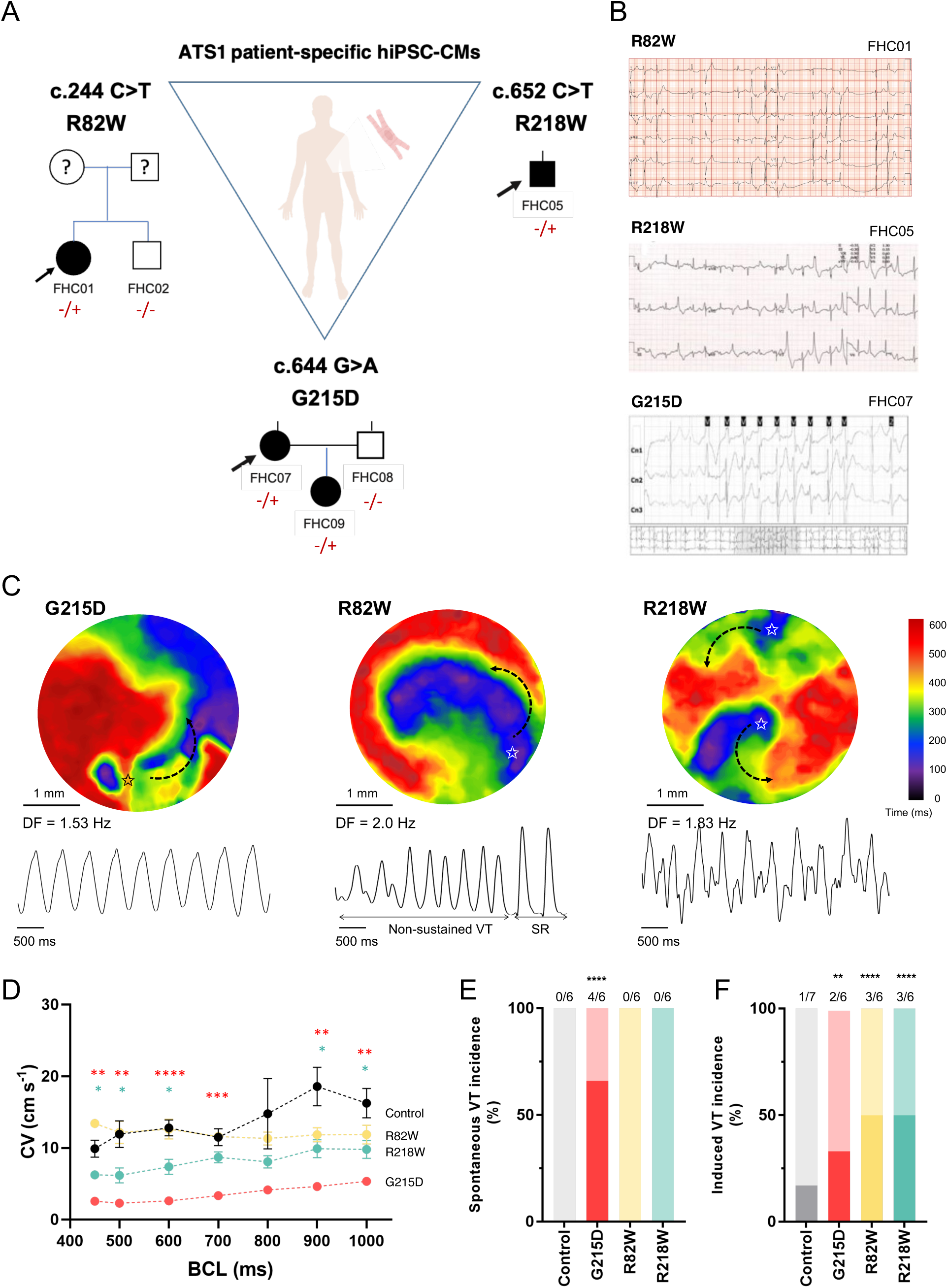
iPSC-CMs from patients carrying Kir2.1-PIP2 mutations served for the risk stratification validation. **A.** Kir2.1-PIP2 variants and ECG cardiac phenotype of three ATS1 subjects. Specific genetics and family history of three probands carrying Kir2.1 variants R82W, R218W and G215D. Males and females are marked with squares and circles, respectively. Mutation carriers are marked with (−/+) and non-carriers with (−/−). **B.** Representative ECG of probands showing a couple of extrasystoles (R82W), ventricular couplets (R218W) and run of VT (G215D). **C.** Representative arrhythmic events of each mutant monolayer group (Kir2.1^G215D^ *left*, Kir2.1^R82W^ *middle* and Kir2.1^R218W^ *right*). Below each map is a single pixel recording revealing varying patterns of monomorphic and polymorphic reentrant tachycardia sustained by one or two rotors. The star indicates the initiation of the reentry. **D.** CV restitution values for patient-specific Kir2.1^G215D^ (red), Kir2.1^R82W^ (yellow) and Kir2.1^R218W^ (green) iPSC-CM monolayers. Kir2.1^G215D^ and Kir2.1^R218W^ were lower at all BCLs (BCL=100, 150, 200, 250 and 300 ms; n=6 monolayers per condition)**. E.** Bar graph of spontaneous VT-like incidence for each monolayer group (n=6 monolayers per group). **F.** Bar graph of induced VT-like incidence for each monolayer group (n=6 monolayers per group). Statistics were conducted using TWO-way ANOVA, ONE-way ANOVA test and Mann-Whitney test, accordingly. Asterisk (*) indicates significance against control (*p<0.05, **p<0.01, ***p<0.001, ****p<0.0001).

Optical mapping experiments evidenced distinct electrophysiological phenotypes among the patients and provided critical insights for proarrhythmic risk assessment. Reentrant arrhythmias with variable patterns were observed in monolayers generated for each of the three Kir2.1-PIP2 variants, as illustrated by the corresponding maps and pseudo-ECGs (**Figure 3C**). CV was significantly slower in Kir2.1^G215D^ and Kir2.1^R218W^ monolayers compared to control across BCL raging from 400 to 1000 ms (**Figure 3D**). At a BCL of 1000 ms, CV in Kir2.1^G215D^ monolayers was approximately 30% slower than in controls. Notably, there was a remarkable gradation in CV across all groups: Control > R82W > R218W > G215D. Importantly, these CV differences aligned with the differences in the occurrence of spontaneous arrhythmias. While reentrant arrhythmias could be induced in all three mutant groups, spontaneous arrhythmias were observed only in Kir2.1^G215D^ monolayers, likely reflecting their abnormally low CV (**Figure 3E-F**).

### Conduction velocity is reduced in ATS1 mutant mouse hearts

To further validate the electrophysiological findings observed in patient-derived cells, we generated mouse models carrying the same Kir2.1 variants (G215D, R82W, R218W). Mice received wild-type or mutant Kir2.1 channels via cardiac-specific, intravenous AAV9-mediated delivery, as previously described (**Supplementary Figure 3**).^14,32^ Cardiac-specific expression was confirmed by tdTomato fluorescence 8 weeks post-AAV transduction, which was absent in non-infected hearts (**Supplementary Figure 3B**). Immunohistochemistry showed ∼95% global ventricular AAV9 infection (**Supplementary Figure 3D-E**), as reported previously,^14,29,33,34^ with ∼50% of cardiomyocytes expressing at least three viral genomes per cell. ATS1 mouse hearts showed normal structure, as assessed by H&E staining (**Supplementary Figures 3C**) and echocardiography (**Supplementary Figure 4**).

Optical mapping in isolated, Langendorff-perfused hearts of mice carrying mutations that affect Kir2.1-PIP2 interactions enabled measurements of ventricular epicardial CV across five different BCLs (100 to 300 ms). CV was reduced at all cycle lengths in all three of mutant hearts compared to controls (**Figure 4A and B**). However, the magnitude of reduction in CV differed among variants (**Figure 4A**). In **Figure 4B**, superimposed CV restitution curves revealed a gradation consistent with that observed in patient-specific iPSC-CMs monolayers (Control > R82W > R218W > G215D). Kir2.1^G215D^ mutant hearts were the most severely affected, exhibiting a ∼2.2-fold CV reduction compared with Kir2.1^WT^ (at BCL=300ms, Kir2.1^G215D^ 19.5 ± 1.4 cm s^-1^ *vs* Kir2.1^WT^ 43.7 ± 3.3 cm s^-1^, p<0.0001). Kir2.1^R218W^ hearts presented an intermediate CV reduction (at BCL=300 ms, Kir2.1^R218W^ 28.3 ± 3.3 cm s^-1^, p<0.0001), whereas Kir2.1^R82W^ displayed only a modest trend toward reduced reduced CV (**Figure 4B**). In contrast, APD did not show significant differences among mutant groups (**Figure 4C**). Overall, these results demonstrate that *KCNJ2* G215D mutation creates a markedly dysfunctional electrical substrate, conferring a greater susceptibility to both spontaneous and inducible life-threatening ventricular arrhythmias compared to the R82W and R218W variants.

**Figure 4.**
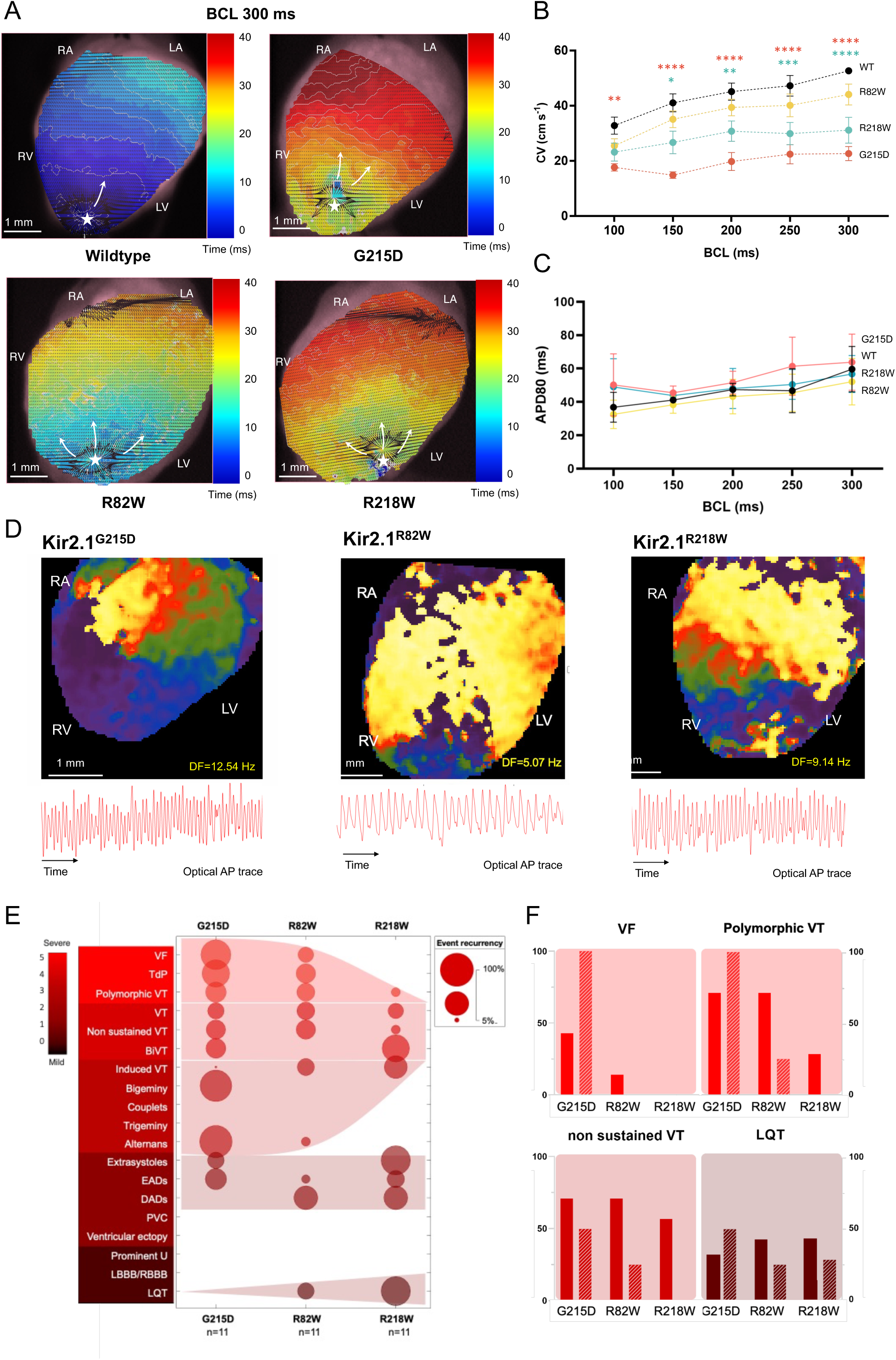
ATS1 mutant mice CV is reduced differently in each of the three mutant heart groups and show reentrant arrhythmias with varying rotor patterns. **A.** Representative velocity maps with local vectors and activation isochrones (2 ms/isochrone) of WT and ATS1 mutant hearts paced at BCL=300 ms. Pacing site is marked with *. **B.** CV restitution curves paced at distinct BCLs (BCL=100, 150, 200, 250 and 300 ms; N=7 animals per condition; Kir2.1^WT^ in black, Kir2.1^G215D^ in red, Kir2.1^R82W^ in yellow and Kir2.1^R218W^ in green). **C.** AP duration restitution curves paced at distinct basic cycle length (BCLs, N=7 animals per condition). **D.** Reentrant arrhythmias in each ATS1 mutant model. *Top*, snapshot of a phase mapping movie. *Bottom*, single-pixel optical AP trace over time. Example of exceedingly rapid polymorphic VT in Kir2.1^G215D^ (*left*), short run of VT in a Kir2.1^R82W^ mouse (*middle*), and TdP-like polymorphic VT in Kir2.1^R218W^ (*right*). **E.** Bubble plot showing the incidence of arrhythmic events in three different ATS1 mice hearts that recaps the stratification of ATS1 patients based on cardiac arrhythmia risk (N=11 animals per condition). Cardiac events are ordered according to severity and cardiac score-risk (5 = severe in red, 0 = mild in brown). **F.** Comparison of incidence of cardiac events between mice (solid) and human (dashed) datasets of Kir2.1-PIP2 mutants, displayed as percentage of frequency of the events (%). Statistical analyses were conducted using TWO-way ANOVA, followed by Tukey’s multiple comparisons. Asterisk (*) indicates statistical differences against WT (*p<0.05, **p<0.01, ***p<0.001, ****p<0.0001).

### Optical mapping enables validation of ATS1 arrhythmogenic stratification

Optical mapping, combined with pseudo-ECGs, allowed us to identify and quantify the types of spontaneous arrhythmias occurring in the hearts of mice carrying mutations that affect Kir2.1-PIP2 interactions. All three mutant groups exhibited a range of sustained and non-sustained ventricular reentry with variable duration and incidence (**Figure 4D**). Classification of these arrhythmias (**Figure 4E**) revealed that Kir2.1^G215D^ hearts gad the highest incidence of VT (e.g., TdP and polymorphic VT) and VF episodes. In contrast, Kir2.1^R82W^ and Kir2.1^R218W^ hearts displayed milder arrhythmic phenotypes, such as bidirectional VT, extrasystoles, and long QT (LQT)–like abnormalities. WT hearts did not show any arrhythmic events, as evidenced by the preserved CVs of 35 to 55 cm/s at BCLs ranging between 100 and 300 ms (see **Figures 4A and B**). Interestingly, VF, polymorphic VT, non-sustained VT, and LQT emerged as the key arrhythmic features that most accurately recapitulate the ATS1 phenotypes across both human and mouse datasets (**Figure 4F**). These findings support the validity of the cardiac-specific AAV9-mediated mouse models to explore mutation-specific mechanisms of ventricular arrhythmias and SCD.

### In silico modelling and molecular dynamics reveal ATS1 mutation-specific atomic mechanisms

We subsequently performed in silico experiments to investigate the atomic-level structural rearrangements and functional alterations caused by mutations that affect Kir2.1-PIP2 interactions. Homology modeling showed that the COOH and NH2 terminals are mutation hotspots (66.7% and 28.2%, **Figure 5A**), with fewer mutations in the transmembrane region (5.1%) and channel pore lining (10.3%). Severe mutations affecting Kir2.1-PIP2 interactions are closer to the channel pore (R=0.2, p=0.21; **Figure 5B**), with these functionally severe mutations averaging 12.02 ± 3.87 Å from the pore versus greater distances for less severe mutations (moderately severe: 17.55 ± 2.98 Å; moderate: 17.58 ± 3.35 Å; mild: 16.07 ± 3.08 Å; **Figure 5B**). Mutations G215D, R82W and R218W reside in the center and intermediate lining of the protein structure. To further assess their functional impact, we conducted atomic resolution molecular dynamics (MD) simulations (2000 ns) for each ATS1 variant. All three mutations impaired PIP2 binding to varying degrees at key residues (R^80^W^81^R^82^ and K^182^–-K^185^–K^187^K^188^ motifs), with greater disruption observed in heterotetramers compared to homotetramers. Overall, PIP2 interactions were most severely disrupted by R82W, followed by G215D, and least affected by R218W (**Figure 5C-D**).

**Figure 5.**
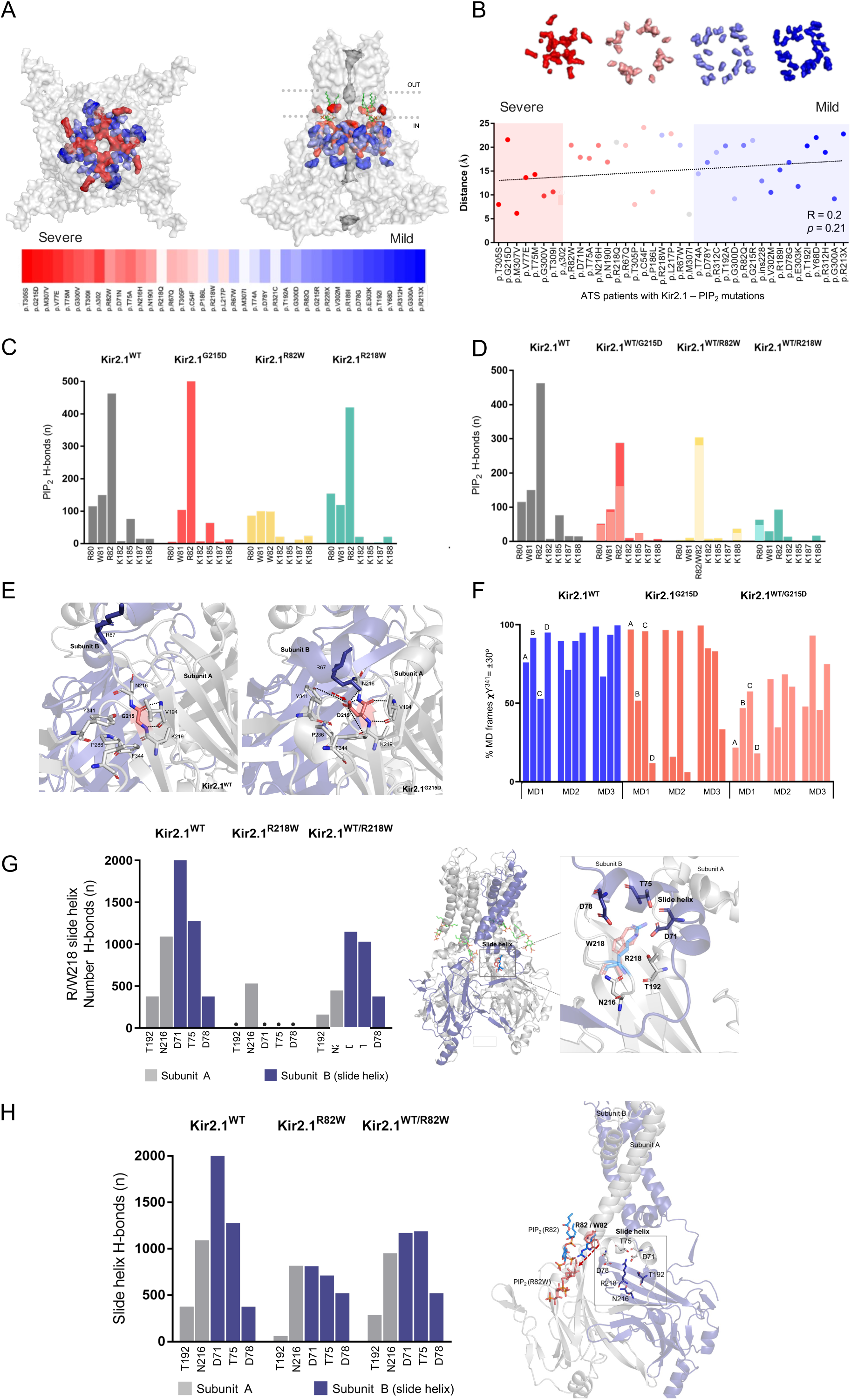
Local association and Kir2.1-PIP2 interactions are impaired ATS1 mutations. **A.** Top-view (left) and frontal view (right) of Kir2.1 protein structure. The more severe mutations based in the stratification are highlighted in red, while less severe mutations are in blue, showing their distribution in the bottom panel. Green sticks represent PIP2 at its binding location in the transmembrane region (N=39 mutations). **B.** Scatter plot displaying distances of mutations to the channel pore (N=39; R=0.2, p=0.21), shaded sections correlate with colored regions in A. Statistical analysis was performed using Pearson correlation. **C-D.** Number of h-bonds of homotetrameric (C) and heterotetrameric (D) mutants (Kir2.1^WT^ in black, Kir2.1^G215D^ in red, Kir2.1^R82W^ in yellow and Kir2.1^R218W^ in green) presented in Kir2.1 residues (R80W81R82 and K182--K185-K187K188 motifs) directly interacting with PIP2**. E.** Local environment of residue G215 in Kir2.1^WT^ (top), and Interactions of D215 in Kir2.1^G215D^ with surrounding amino acids (bottom). Red arrows indicate new G215D-Y314 interaction. Black dashed lines indicate h-bonds. **F.** Shift of first Y341 c dihedral. Bar plot representing the percentage of frames where Y341 dihedral is maintained within ±30o of initial position. A, B, C, D indicates each subunit conforming the Kir2.1 tetramer (Kir2.1^WT^ in blue, homotetrameric Kir2.1^G215D^ in red, and heterotetrameric 2:2 Kir2.1^G215D^ in pink). Mean values of 3 MD replicas per condition. **G.** Quantification of total number of h- bonds in the R/W218 network in mutant Kir2.1^R218W^ for 2000 ns MD (subunit A with T192, N216 in gray, neighboring subunit with slide helix (D71, T75, D78) in purple (*left*). Detailed view of relevant residues involved in the R/W218 mutant network, visualized with sticks (*right*). **H.** Quantification of total number of h- bonds in the R/W218 network in Kir2.1^R82W^ for 2 μs MD (subunit A with T192, N216 in gray, neighboring subunit with slide helix (D71, T75, D78) in purple (*right*). Graphical representation of R/W218 localization within Kir2.1^WT^ and structure Kir2.1^R82W^ (*left*). Mean values of 3 replicates per condition.

Detailed analysis revealed distinct mutation-specific mechanisms. G215D induced a conformational change at the dimerization interface (Y341), likely altering channel assembly or biophysical properties.^35,36^ While G215D preserved PIP2 interactions better than R82W, its greater disruption of the dimerization interface rendered it the most functionally severe variant. Modulation of the Y341 sidechain dihedral angle during MD simulation partially reduced, but did not fully abolish, channel activity (**Figure 5E-F**). In contrast, R82W and R218W mainly disrupted the slide helix, with R218W causing the most pronounced loss of the wildtype hydrogen bond network (**Figure 5G-H**).

## Discussion

Our pooled, patient-level analysis comprised data from 225 individuals harboring Kir2.1 protein mutants that disrupt Kir2.1-PIP2 interactions. We focused on PIP2 mutations, as they account for over 50% of ATS1 variants.^18^ Despite this shared mechanism, a broad spectrum of mutation-dependent phenotypes was observed across patients. ATS1 mutations were stratified into three distinct clusters: Cluster #1 included mutations associated with the highest incidence of arrhythmic events; Cluster #2 encompassed mutations producing mild and heterogeneous electrical phenotypes; and Cluster #3 comprised mutations with minimal cardiac electrical manifestations. Regression analysis identified variables (p<0.01) strongly associated with increased susceptibility to cardiac arrhythmias, including, in order of ranking, non-sustained VT, SCD, bigeminy, and syncope. Certain Kir2.1 mutations (e.g., p.T305S, p.G215D, and p.M307V) were significantly more arrhythmogenic than others (e.g., p.Y68D, p.R321H, and p.G300A), highlighting a clear mutation-dependent hierarchy of severity.

Pharmacologic treatment of ATS1 is not yet based on a detailed mechanistic understanding of the syndrome, and the possibility that some mutations may be more malignant than others is often overlooked. This likely explains why therapeutic success derived from the empirical use of beta-blockers and class I antiarrhythmics is limited.^30,37^ ICD implantation is recommended to prevent SCD, but it can be ethically complex, especially in young patients, and is not always fully effective.^7,38^ Stratification of ATS1 mutations affecting Kir2.1-PIP2 interactions (over 51% of loss-of-function variants) shows that some mutations are more arrhythmogenic than others.^4,18,39,40^ Studies demonstrate a spectrum of arrhythmic risk depending on the mutation. Other mutations, such as those impacting channel trafficking or conductance, also vary in severity. For example, the trafficking-defective Δ314-315 mutation traps Kir2.1 and Na_V_1.5 at the Golgi, causing high arrhythmia risk.^14^ In contrast, the C122Y mutation disrupts disulfide bonds in Kir2.1, reducing membrane expression by a different mechanism.^29^

iPSC-CMs derived from ATS1 patients with Kir2.1-PIP2 mutations demonstrated distinct effects of each variant on arrhythmias and cardiac conduction. The severity of arrhythmias differed among patients, and these variations were mirrored in iPSC-CM monolayers. CV was variably reduced (Control > R82W > R218W > G215D), with the Kir2.1^G215D^ variant conferring the highest risk for life-threatening arrhythmias. These findings underscore the need for personalized treatment strategies for ATS1 patients, as different Kir2.1 mutations result in unique mechanisms and phenotypes.

AAV9 technology^33^ enabled the generation of ATS1 mouse models^14,29,33,34^ using gene constructs containing mutations Kir2.1^G215D^, Kir2.1^R82W^, and Kir2.1^R218W^. These were compared with a Kir2.1^WT^ mice serving as a control to validate the in vitro human models. Although these mutations have previously been studied in heterologous expression systems,^41–43^ such approaches cannot replicate the complex molecular cardiac environment that may conceal differences among mutants. This study represents the first introduction of these mutations into a mouse model, allowing in vivo assessment of the arrhythmogenic mechanisms associated with Kir2.1-PIP2 impairment in ATS1. Notably, AAV9-mediated delivery of human WT or mutant Kir2.1 sequences neither altered endogenous Kir2.1 expression nor induced Kir2.1 overexpression in the heart, successfully establishing a heterozygous condition with a dominant negative effect, mirroring the genetic background of affected patients.^14,29^

Cardiac-specific ATS1 mouse models harboring Kir2.1-PIP2 mutations faithfully replicated the arrhythmia phenotypes observed in patients, including ventricular arrhythmias. Ex vivo optical mapping further validated that these models accurately recapitulate the ATS1 phenotype and provide a robust framework for proarrhythmic risk assessment. Among the mutations, Kir2.1^G215D^ exhibited the highest incidence of arrhythmic events, including VF, followed by Kir2.1^R82W^ and Kir2.1^R218W^, supporting the proposed risk stratification. Notably, this study is the first to investigate mutation-specific arrhythmia severity in ATS1, revealing a spectrum of arrhythmia risk linked to varying conduction velocities (Control > R82W > R218W > G215D). While mutations such as R82W may still permit flecainide therapy, more severe mutations that significantly reduce I_K1_ and sodium channel function should be managed with alternative approaches.^29^

This research reveals a key link between mutation location and symptom severity in dysfunctional Kir2.1-PIP2 interactions. The COOH and NH2 terminals are major hotspots. Most severe cardiac mutations cluster near the channel pore, supporting earlier studies on LQT1^44^ and LQT2^45,46^ and highlighting the link between domain-specific mutations and arrhythmic risk. Kir2.1-PIP2 disrupting variants must reach the sarcolemma before causing pathophysiological changes.^47^ MD simulations using the pre-opened channel state bound to PIP2 examined three Kir2.1-PIP2 mutations (Kir2.1^G215D^, Kir2.1^R218W^, Kir2.1^R82W^), spanning ATS1 cardiac arrhythmia severity. For milder mutants (Kir2.1^R218W^, Kir2.1^R82W)^, loss of PIP2 interaction—especially at R82, a key anchoring residue^16^—likely drives pathogenicity. In Kir2.1^G215D^, disruption of the dimerization interface^48^ and effects on the Y341 residue^36^ may explain its greater severity. Although R218W affects a secondary PIP2 binding hotspot, it does not cause a severe cardiac phenotype. Overall, MD simulations suggest that more profound protein structure changes, not just PIP2 impairment, are needed for severe ATS. This type of in silico studies could help the discovery of new drugs useful for arrhythmia treatment.^49^ As ATS1 is not explained solely by I_K1_ differences, suggesting additional modifiers. Although few new antiarrhythmic drugs are in development, SGLT2 inhibitors like dapagliflozin and empagliflozin have been suggested to enhance I_Na_ and I_K1_, thereby restoring cardiac excitability.^50^ Overexpressing *SNTA1* can also increase *I_K1_* and *I_Na_*, offering antiarrhythmic effects in certain muscular dystrophy patient cells.^27^ These findings suggest that future treatments—including gene therapies that raise Kir2.1 and Na_V_1.5 levels—could provide new, targeted options for ATS1 arrhythmia prevention.^51^

## Limitations

ATS1 is a rare disease, affecting about 1 in a million people.^8,52^ The small number and limited size of studies, combined with inconsistent findings, complicate meta-analyses and risk assessment. No patient-level pooled analyses currently exist for rare diseases like ATS1.^53^ This non-randomized study of 225 ATS1 patients adhered to PRISMA guidelines, though a larger validation cohort would have strengthened the conclusions. Despite these challenges, four different scoring methods—with distinct quantitative approaches—showed broad agreement in classifying mutations. The ATS1 cardiac risk score was externally validated in candidates who underwent the proposed stratification. However, only three mutations from cluster #1 were experimentally evaluated; broader molecular studies encompassing additional clusters are necessary. In-vitro, ex-vivo, and in-silico analyses demonstrated that mutations disrupting Kir2.1-PIP2 interactions increase susceptibility to arrhythmias and affect the risk score. MD studies also had limitations, including the use of mixed Kir2.x isoform sequences and relatively short (2000 ns) simulations, which require a fixed system composition (e.g., membrane, water content) to be set in advance.^54^

## Clinical Perspective

These results refine the understanding of the electrical abnormalities identified in ATS1 patients and provide mechanistic insight into the arrhythmogenic consequences of *KCNJ2* variants that disrupt Kir2.1–PIP2 interactions. Importantly, our findings demonstrate that variability in arrhythmic risk is driven by mutation-specific effects on channel regulation and ventricular conduction, supporting a stratified model in which distinct molecular mechanisms underlie discrete clinical risk profiles.

## Data Availability

The data supporting the findings of this study are available within the article and its Supplementary Materials. Additional data underlying the results presented in this manuscript are available from the corresponding author upon reasonable request

## Acknowledgements

We thank and acknowledge the contribution of Dr. María Luisa Peña, Dr. Helena Llamas (Hospital Virgen del Rocío), Dr. Jose María Larranaga (Hospital A Coruña) Dr. Marina Martínez-Moreno (Hospital General Universitario de Elche), Dr. Juan Kasaki (Great Ormond Street Hospital), and Dr. David Calvo (Hospital Universitario Central de Asturias) for contributing with their own patients and build the validation cohort. We thank the Centro de Supercomputación de Galicia (CESGA) for use of the Finis Terrae III supercomputer to perform molecular dynamics studies. We thank Dr. Stuart Pocock (London School of Hygiene and Tropical Medicine) and Manuel Vazquez-Sanchez (NYU Grossman School of Medicine) for reviewing the statistical analysis, and Dr. Carlos Morillo for reviewing the manuscript and their insightful comments in the preparation of this work.

## Abbreviations

ATS1: Andersen-Tawil syndrome type 1
AP: Action potential
CV: Conduction velocity
ICD: Implantable cardioverter defibrillator
iPSC-CMs: Induced pluripotent stem cell-derived cardiomyocytes
MD: Molecular dynamics
PIP2: Phosphatidylinositol-4,5-biphosphate
SCD: Sudden cardiac death
VF: Ventricular fibrillation
VT: Ventricular tachycardia

